# Association between Hemoglobin Dynamic Trajectories and 28-Day Mortality in Elderly Patients with Sepsis: A retrospective cohort study

**DOI:** 10.1101/2025.06.17.25329817

**Authors:** Yan Zeng, Jun Wang, Caoyi Liu, Jingwei Zhang

**Affiliations:** Department of Hematology, Chengdu Second People’s Hospital, Chendu, China; Department of BIood Transfusion, Chengdu Second People’s Hospital, Chengdu, China

**Keywords:** Elderly Patients, Sepsis, Mortality, Joint latent class model (JLCM)

## Abstract

**Background:** Anemia is a common complication in sepsis and is associated with poor prognosis. However, there are few studies on the dynamic changes in hemoglobin (Hb) levels and their relationship with prognosis in elderly patients with sepsis. This study aims to retrospectively investigate the relationship between Hb trajectories and 28-day mortality in this population.

**Methods:** This retrospective observational study used data from 4,961 elderly patients with sepsis from the MIMIC-IV database. A joint latent class model (JLCM) was employed to analyze Hb trajectories. Time-dependent piecewise Cox regression models and Kaplan-Meier survival curves were used to assess the relationship between trajectories and 28-day mortality. Sensitivity analyses included the Schoenfeld residual test, subgroup analysis, and E-value assessment.

**Results:** Hb trajectories were classified into three patterns: Class 1 with persistently low levels, Class 2 with medium levels and a gradual decline, and Class 3 with a rapid decline from higher levels. Using Class 2 as the reference, multivariable Cox regression showed that Class 1 (HR = 1.38, 95% CI = 1.18–1.62, p < 0.001) and Class 3 (HR = 1.21, 95% CI = 1.03–1.42, p = 0.020) were associated with higher 28-day mortality. Kaplan-Meier curves indicated the lowest survival probability in Class 1 and the highest in Class 2. Gender-specific analysis revealed consistent trajectory patterns between males and females, but higher Hb values in males. The predictive power of the rapid decline pattern for early mortality was stronger in males.

**Conclusion:** The Hb trajectories in elderly sepsis patients are significantly associated with early mortality. Persistently low levels and rapid declines in Hb are indicators of poor prognosis.

## Introduction

Sepsis, a systemic inflammatory response syndrome induced by infection, represents one of the most prevalent and lethal conditions encountered in the Intensive Care Unit (ICU), marked by significant morbidity and mortality rate[1]. Although advancements in the early detection and therapeutic approaches for sepsis have been made in recent years, the mortality rate among sepsis patients remains alarmingly high. Key objectives in the management of sepsis within the ICU include the early evaluation of patient prognosis, prompt identification of high-risk individuals, and the implementation of effective interventions [2]. Anemia is a frequently observed complication in patients with sepsis, with an incidence rate of 58% during ICU hospitalization. Among critically ill patients whose ICU stay extends beyond seven days, 71.8% experience varying degrees[3,4]

Hemoglobin (Hb) is the most frequently utilized biomarker for evaluating the presence and severity of anemia. It not only serves as an indicator of a patient’s hematological status but is also associated with the severity of sepsis, multiple organ failure, and adverse prognostic outcomes [5,6]. Nonetheless, prior research has predominantly concentrated on the baseline values of static hemoglobin levels, thereby neglecting the dynamic variations of hemoglobin during sepsis treatment. These dynamic changes in hemoglobin, akin to platelet parameters, may provide a more precise reflection of the clinical progression and mortality risk in sepsis patients, as evidenced by their acute decline, short-term fluctuations, or long-term stability [7–9]. Current studies have indicated that in populations without traumatic brain injuries, dynamic changes in hemoglobin levels correlate with mortality in patients who do not have sepsis. However, this correlation is not observed in those with sepsis, suggesting that the relationship between dynamic hemoglobin trajectories and prognosis varies among populations with different underlying diseases and complications[10]. The incidence and mortality rates of sepsis significantly increase with age in the elderly[11]. Elderly individuals at high risk, due to factors such as frailty, complex underlying diseases, long-term chronic anemia, or malnutrition, may exhibit distinct hematological characteristics compared to the general population [12]. Nevertheless, there remains a lack of systematic research examining the correlation between hemoglobin change trajectories and early mortality in elderly patients with sepsis. Therefore, exploring the relationship between the dynamic trajectory of hemoglobin changes and the prognosis of elderly patients with sepsis is not only clinically significant but also contributes to enhancing the prognostic evaluation system for sepsis.

This study aims to explore the association between dynamic changes in Hb levels among elderly sepsis patients, based on data from the Medical Information Mart for Intensive Care (MIMIC-IV, version 3.0), and 28-day mortality. The findings provide new insights for early risk assessment in this demographic through a retrospective analysis.

## Materials and Methods

### Population and Data Collection

This study utilized data from MIMIC-IV, version 3.0, which includes information on over 70,000 patients admitted to the ICUs of Beth Israel Deaconess Medical Center in Boston, MA, from 2008 to 2019 [13]. Access to the database required completion of the “CITI Data or Specimens Only Research” training course available on the National Institutes of Health website, and approval was granted to extract data for research purposes (Certificate No for Zhang: ID: 65677952). The inclusion criteria were as follows: (1) patients met the diagnostic criteria for sepsis according to the Sepsis-3 guidelines, specifically requiring a suspected infection and a change of at least 2 points in the Sequential Organ Failure Assessment (SOFA) score [9]; (2) patients had baseline platelet count data at the time of admission and at least three platelet count measurements recorded within 28 days of admission; (3) age ≥ 65 years. Patients were excluded from the study if they met any of the following criteria: (1) repeated ICU admissions; (2) an admission duration of less than 72 hours.

Structured Query Language (SQL) with PostgreSQL (version 13.0) and Navicat software (version 17.0) were employed to identify the cohort and extract pertinent clinical data [14]. Demographic information, comorbidities, and laboratory indicators were sourced from the MIMIC-IV database. The platelet laboratory data encompassed measurements taken on days 1, 3, 5, and 7 post-admission to the ICU, as well as the averages for weeks 2, 3, and 4, including the minimum, maximum, and average values recorded during hospitalization. In instances where data for days 3, 5, or 7 were absent, values from the nearest available day were utilized to bridge the gaps. Other laboratory tests extracted maximum, minimum, or average values during hospitalization based on their clinical relevance. The primary outcome of the study was 28-day mortality. Variables with missing data exceeding 10% were excluded from the analysis, in accordance with the recommendations of the Strengthening the Reporting of Observational Studies in Epidemiology (STROBE) statement [15]. Multiple imputation was employed to address variables with missing data of less than 10% [16]. The patients ultimately included will be classified according to their hemoglobin trajectory patterns, and a follow-up analysis of their 28-day mortality will be performed.

### Identification of different trajectories

We utilized the R package lcmm (version 2.0.2) to identify subpopulations exhibiting heterogeneous Hb trajectories [8]. A joint latent class model (JLCM)was employed for the longitudinal analysis. Additionally, we implemented a parametric survival model, where the baseline hazard function adhered to a class-specific Weibull distribution [8]. The optimal number of classes was established by minimizing the log-likelihood, alongside evaluating the Akaike Information Criterion (AIC), Bayesian Information Criterion (BIC), sample-adjusted BIC (SABIC), entropy closest to 1, balanced class proportions, and average posterior probability [10,17]

### Statistical analysis

Continuous variables were represented by means for normally distributed data and by medians for skewed data, with group comparisons conducted using the Kruskal-Wallis test. Categorical variables were expressed as percentages (%), with group comparisons performed using the chi-squared test or Fisher’s exact test. Univariate and multivariate Cox regression analyses were utilized to assess the association between characteristic variables and 28-day mortality. The criteria for adjusting covariates in the multivariate analysis were informed by prior literature, clinical experience, and the outcomes of univariate Cox regression [18]. The covariate selection principles were as follows: (1) After adjustment, if the original hazard ratio (HR) effect of the independent variable on the outcome variable changes by more than 5%; (2) There is no collinearity between the covariates and the independent variable or among the covariates themselves. Collinearity is assessed using the Variance Inflation Factor (VIF), with a VIF of 10 or greater indicating the presence of collinearity. Four models were established using multivariate Cox regression to analyze the association between trajectories and 28-day mortality. The Schoenfeld residual test was employed to verify the assumption of proportional hazards in the Cox analysis[19]. Survival curves were plotted using Kaplan-Meier (KM) analysis, and log-rank tests were conducted. Subgroup analysis and variable regrouping were employed as part of the sensitivity analysis. The E-value for sensitivity analyses and subgroup analysis was utilized to assess the robustness of the observed associations [20]. All analyses were conducted using R Statistical Software (Version 4.2.2, available at http://www.R-project.org, The R Foundation) and the Free Statistics analysis platform (Version 2.1.1, Beijing, China). A two-tailed test was performed, with a significance level set at p < 0.05 [21].

## Results

### Baseline characteristics

This study included a total of 4,961 participants, who were divided into two groups based on whether death occurred within 28 days: the survival group (n = 3,974) and the death group (n = 987). The process of participant inclusion, following the established inclusion and exclusion criteria, is illustrated in S1 Fig. A comparison of the basic characteristics and clinical indicators among the study subjects is presented in Table 1.

**Table 1.**
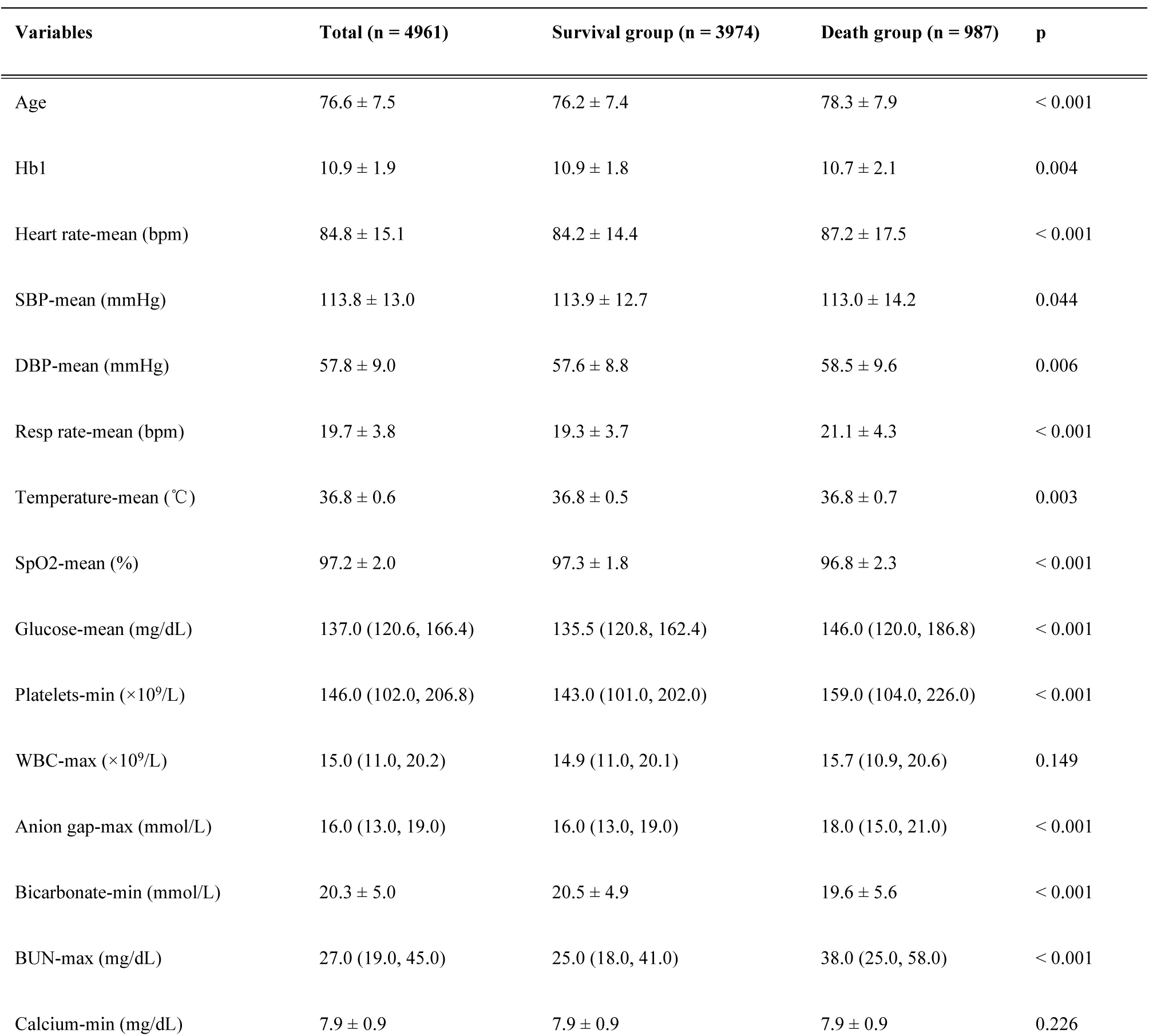
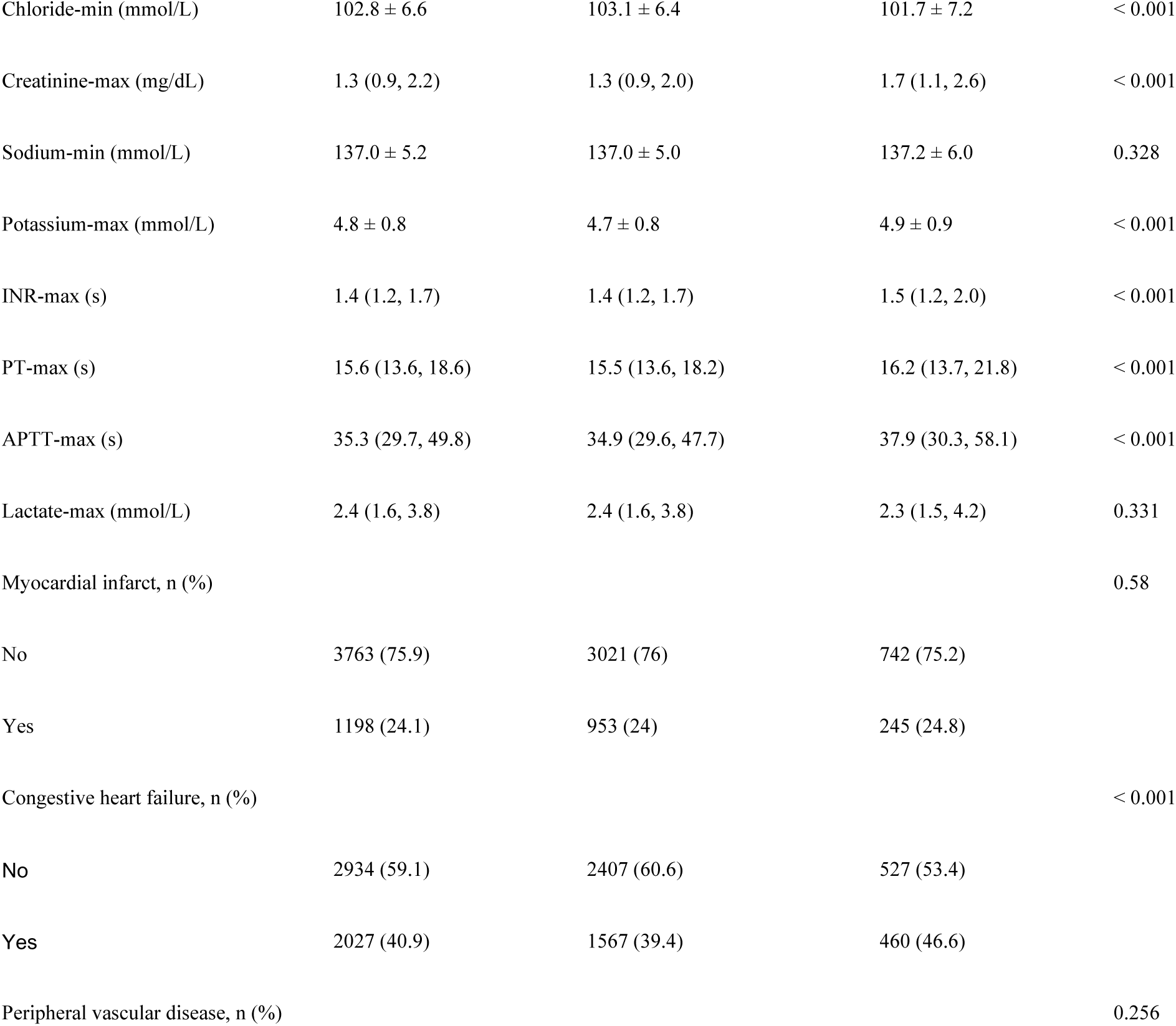
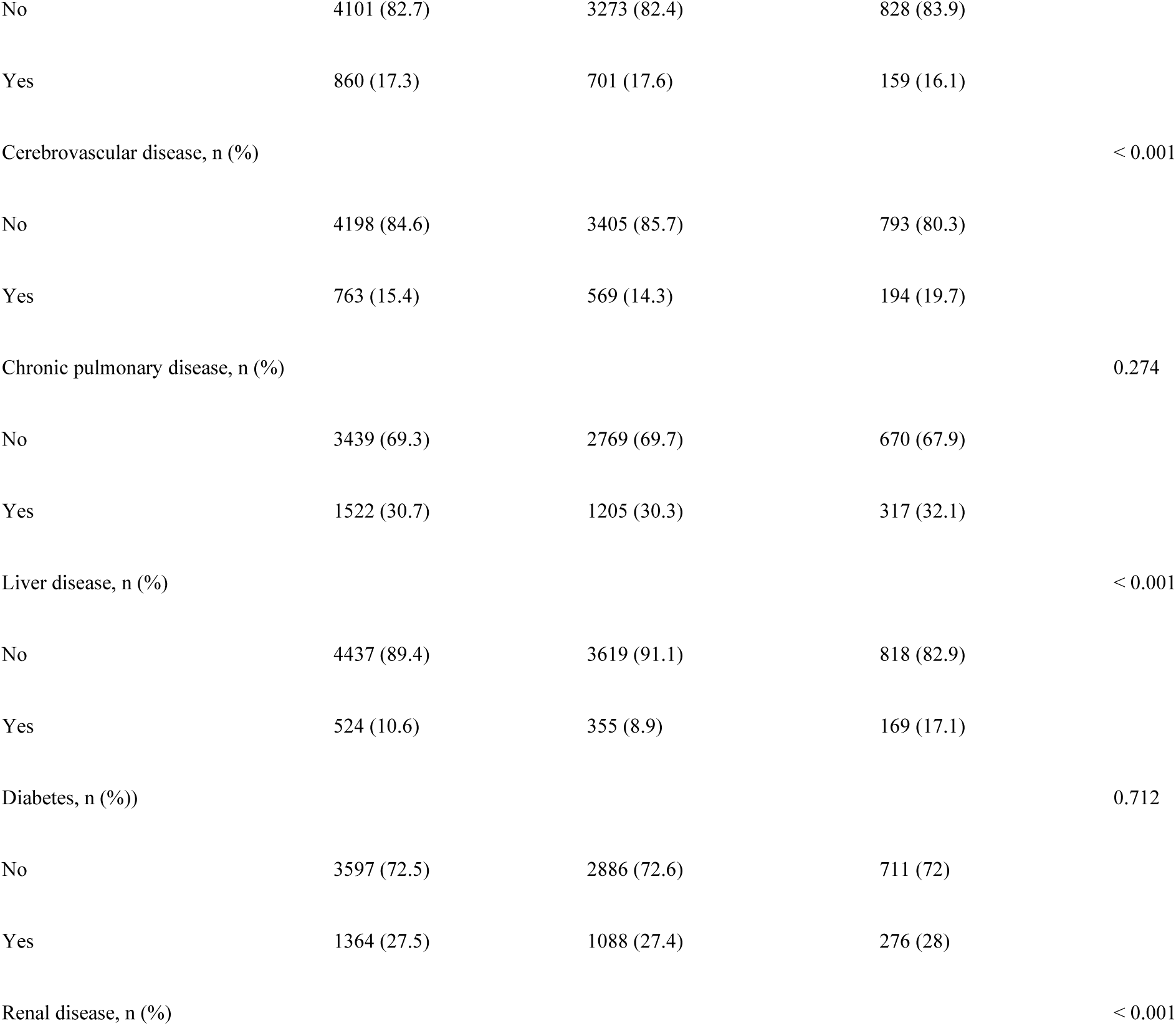
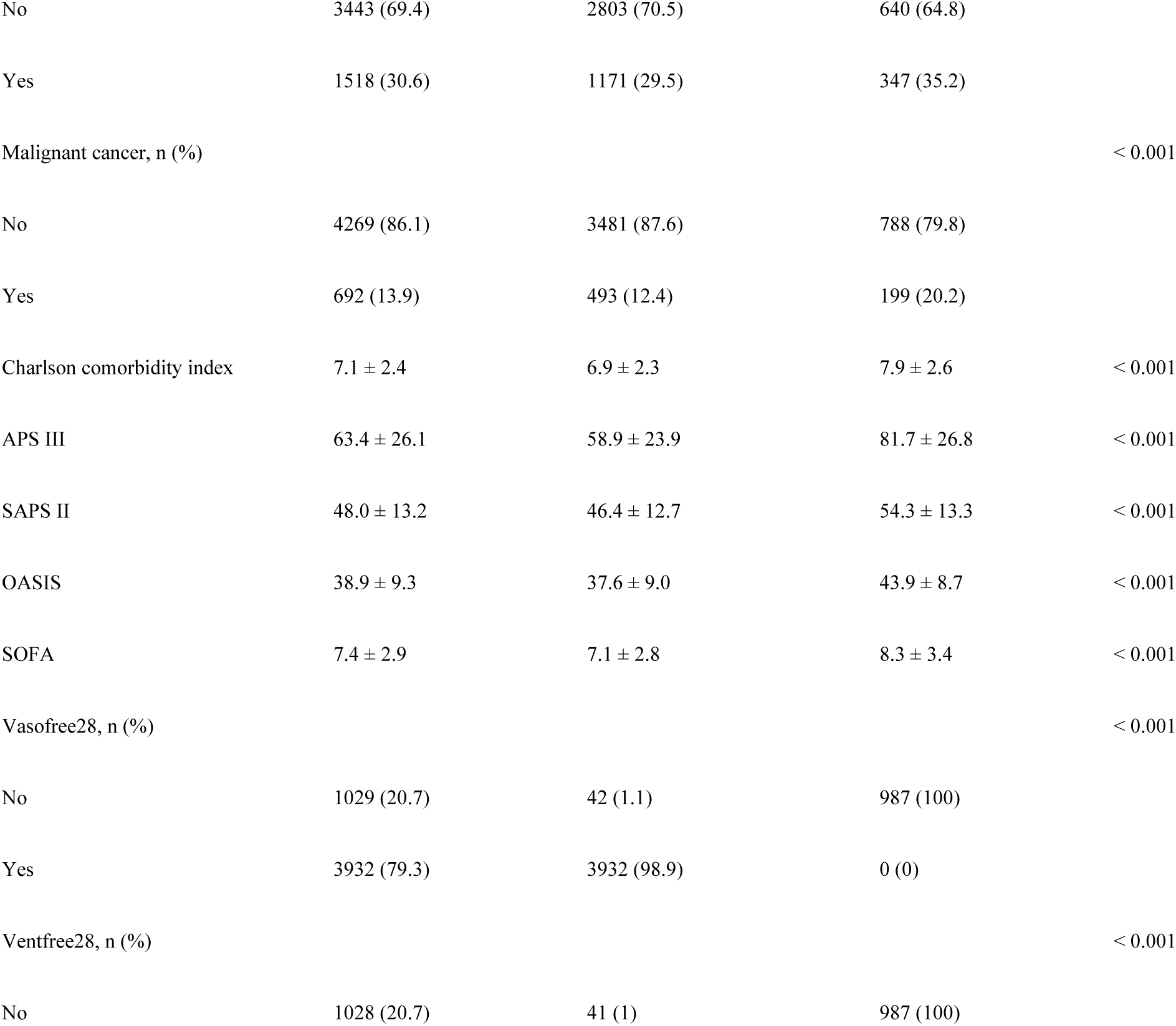
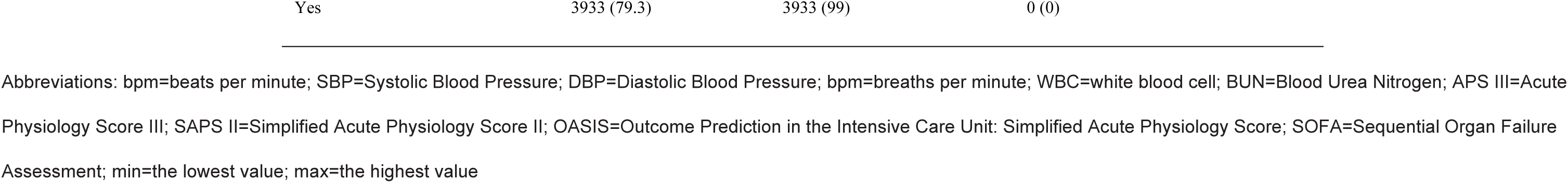
Baseline characteristics of the study population.

Compared to the survival group, patients in the death group were significantly older (78.3 ± 7.9 years vs. 76.2 ± 7.4 years, p < 0.001) and exhibited significantly higher severity of illness, as indicated by APACHE III, SAPS II, and SOFA scores. Multiple blood laboratory indicators (such as white blood cell count, platelet count, urea nitrogen, creatinine, and lactate) as well as physiological indicators (including heart rate, blood pressure, oxygen saturation, and blood glucose) demonstrated significant differences between the two groups, indicating a poorer health status in the deceased group. Furthermore, the prevalence of heart failure, cerebrovascular disease, and liver and kidney diseases was higher in the deceased group (all p < 0.01), and the Charlson Comorbidity Index was also elevated in this group, revealing a greater burden and complexity of diseases.

### Identification of subpopulations using the JLCM

The comparison of different potential trajectory models (2-5 classes) regarding goodness-of-fit and classification effectiveness is presented in Table 2. The 4-class model exhibits the best performance based on the Akaike Information Criterion (AIC) at 73305.1, the Bayesian Information Criterion (BIC) at 73422.3, and the Sample-Size Adjusted BIC (SBIC) at 73365.1. Additionally, it achieves a higher entropy value of 0.53897, which indicates a clearer classification. However, the proportion of Class 3 (9.63%) is significantly lower than that of the other classes (21.06%, 37.92%, and 30.84%). Although the 3-class model is slightly less effective than the 4-class model in terms of AIC (73398.0), BIC (73489.1), SBIC (73444.6), and entropy (0.50263), it still demonstrates clear classification. Furthermore, the proportions of patients in each category of the 3-class model are more balanced (23.63%, 23.48%, and 53.05%), and the probability indicators are higher, suggesting that this model can effectively differentiate between distinct trajectories. In conclusion, the 3-class model is identified as the optimal solution for trajectory analysis. The dynamic trajectories of the three model types are illustrated in Fig 1. The proportions of patients in the four classes are as follows: Class 1 (low-level maintenance, 23.63%); Class 2 (medium-level slow reduction, 53.05%); Class 3 (high-level rapid reduction, 23.48%).

**Fig. 1.**
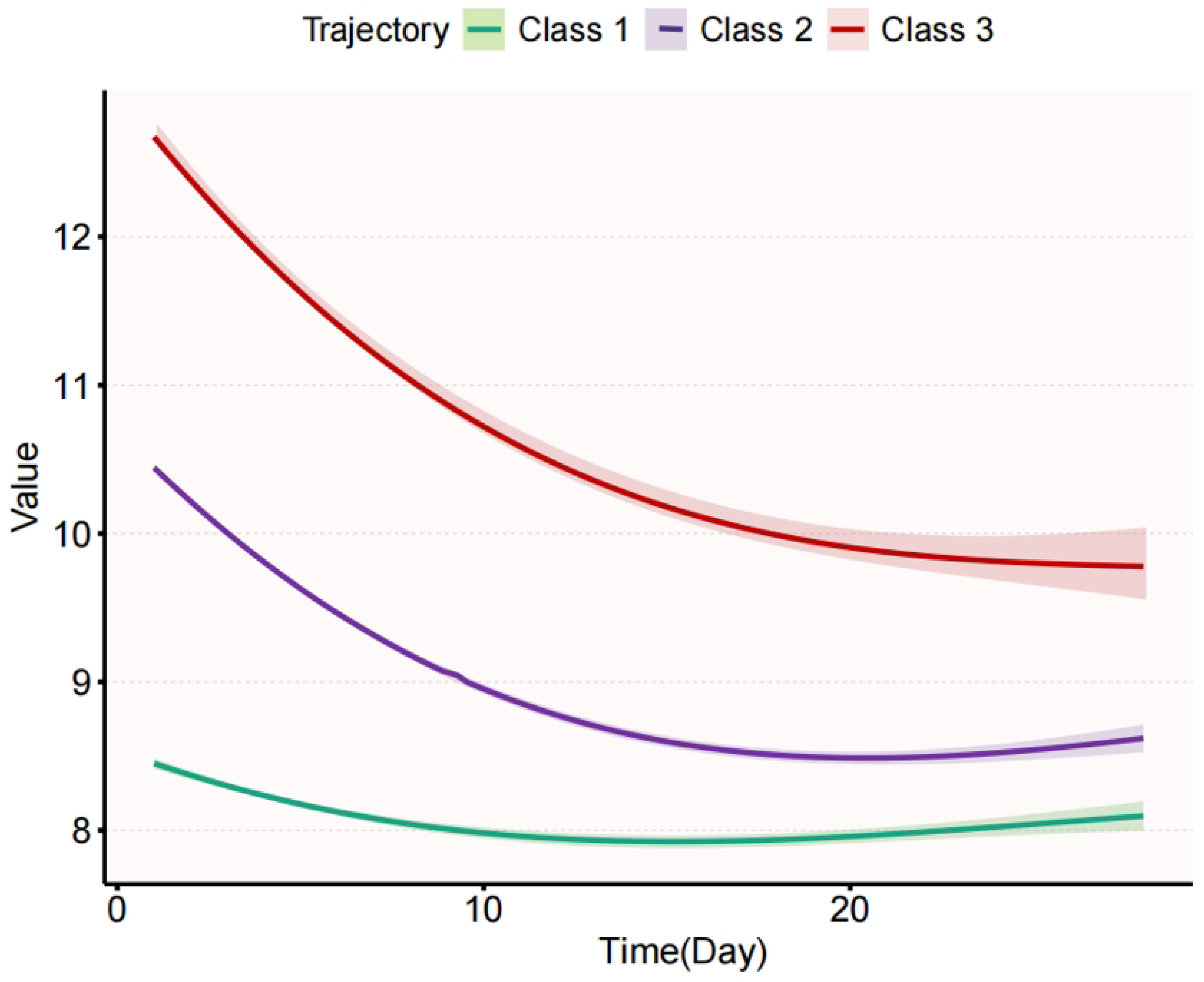
Trajectory curve

**Table 2.** Metrics for determining the optimal trajectories.

| Trajectory | Features |  |  |  |  | Proportion of Patients (%) |  |  |  |  |
| --- | --- | --- | --- | --- | --- | --- | --- | --- | --- | --- |
| Class | Loglikelihood | AIC | BIC | SABIC | Entropy | Class1 ( % ) | Class2 ( % ) | Class3 ( % ) | Class4 ( % ) | Class5 ( % ) |
| 2 | -36790.9 | 73601.8 | 73666.8 | 73635.1 | 0.69848 | 89.256 | 10.743 | NA | NA | NA |
| 3 | -36685 | 73398 | 73489.1 | 73444.6 | 0.50263 | 23.463 | 53.053 | 23.483 | NA | NA |
| 4 | -36634.5 | 73305.1 | 73422.3 | 73365.1 | 0.53897 | 21.608 | 37.915 | 9.6351 | 30.84 | NA |
| 5 | -36601.4 | 73246.8 | 73390 | 73320.1 | 0.62447 | 19.935 | 31.203 | 31.263 | 12.618 | 4.9788 |

Abbreviations: AIC=akaike information criterion, BIC=bayesian information criteria, SABIC=sample-adjusted information criteria

### Clinical Characteristics of Different Hemoglobin Trajectories

The clinical characteristics of various Hb trajectory classes are presented in S1 Table. The results indicate significant differences among the classes in terms of blood pressure, respiratory rate, body temperature, blood oxygen levels, blood glucose, hematological parameters, and coagulation indices (p < 0.05). Notable variations in Hb changes were observed across different trajectories. Class 3 exhibited significantly higher Hb levels at all time points compared to the other classes, peaking at 13.4 g/dL on Day 1, but demonstrated a notable downward trend thereafter. Conversely, Class 1 consistently showed lower Hb levels at all time points, reaching a minimum of 7.0 g/dL on Day 5. Class 2 occupied an intermediate position, displaying a gradual and slow decline while remaining relatively stable within a certain range. Overall, Hb level changes in Class 3 were higher but declined rapidly, indicating a worsening degree of anemia, whereas Class 1 maintained a consistently low level, suggesting a persistent anemic state.

S2 Table presents the results of the univariate Cox regression analysis concerning 28-day mortality. Several clinical indicators—including age, blood gas parameters, biochemical markers, and underlying diseases—are significantly associated with the risk of mortality. Among the Hb trajectory classes, Class 1 (HR 1.64) and Class 3 (HR 1.28) notably elevate the risk of death, indicating that dynamic changes in Hb levels are crucial prognostic factors.

The analysis of covariates influencing Hb trajectory, informed by clinical significance and univariate Cox regression results, is presented in S3 Table. The screening outcomes identified several potential covariates associated with changes in Hb levels, including Hb value of the first day, respiratory rate, body temperature, and various blood gas parameters (such as chloride, base excess, and creatinine), as well as the Charlson comorbidity score, APACHE III, and OASIS score. These covariates demonstrated significant correlations with alterations in Hb trajectory (p < 0.05), with VIF values remaining within an acceptable range. As INR and PT exhibited multicollinearity, INR was ultimately chosen as the covariate for further analysis.

### Time-dependent HR from piecewise Cox model and Kaplan-Meier curves analysis

We conducted the Schoenfeld residual test to verify the assumption of proportional hazards in the Cox analysis. This assumption was validated for the endpoints of death from any cause, resulting in a p-value of 0.646. In the multivariable Cox proportional hazards model.

In the multivariable Cox proportional hazards model presented in Table 3, the unadjusted model (Model 1) revealed that, compared to Class 2, Class 1 significantly increased the risk of mortality (HR 1.64, p < 0.001), while Class 3 also exhibited a notable increase in risk (HR 1.28, p = 0.001). Model 2 adjusted for age and sex, and Model 3 further incorporated laboratory indices based on Model 2. Model 4 demonstrated that after adjusting for all covariates: age, respiratory rate mean, temperature mean, anion gap maximum, bicarbonate minimum, blood urea nitrogen maximum, creatinine maximum, international normalized ratio maximum, SAPS II, OASIS, and the Charlson comorbidity index, the Hb trajectory remained an independent prognostic indicator: Class 1 (HR = 1.38, 95% CI = 1.18–1.62, p < 0.001) and Class 3 (HR = 1.21, 95% CI = 1.03–1.42, p = 0.020), as illustrated in Table 3.

**Table 3.**
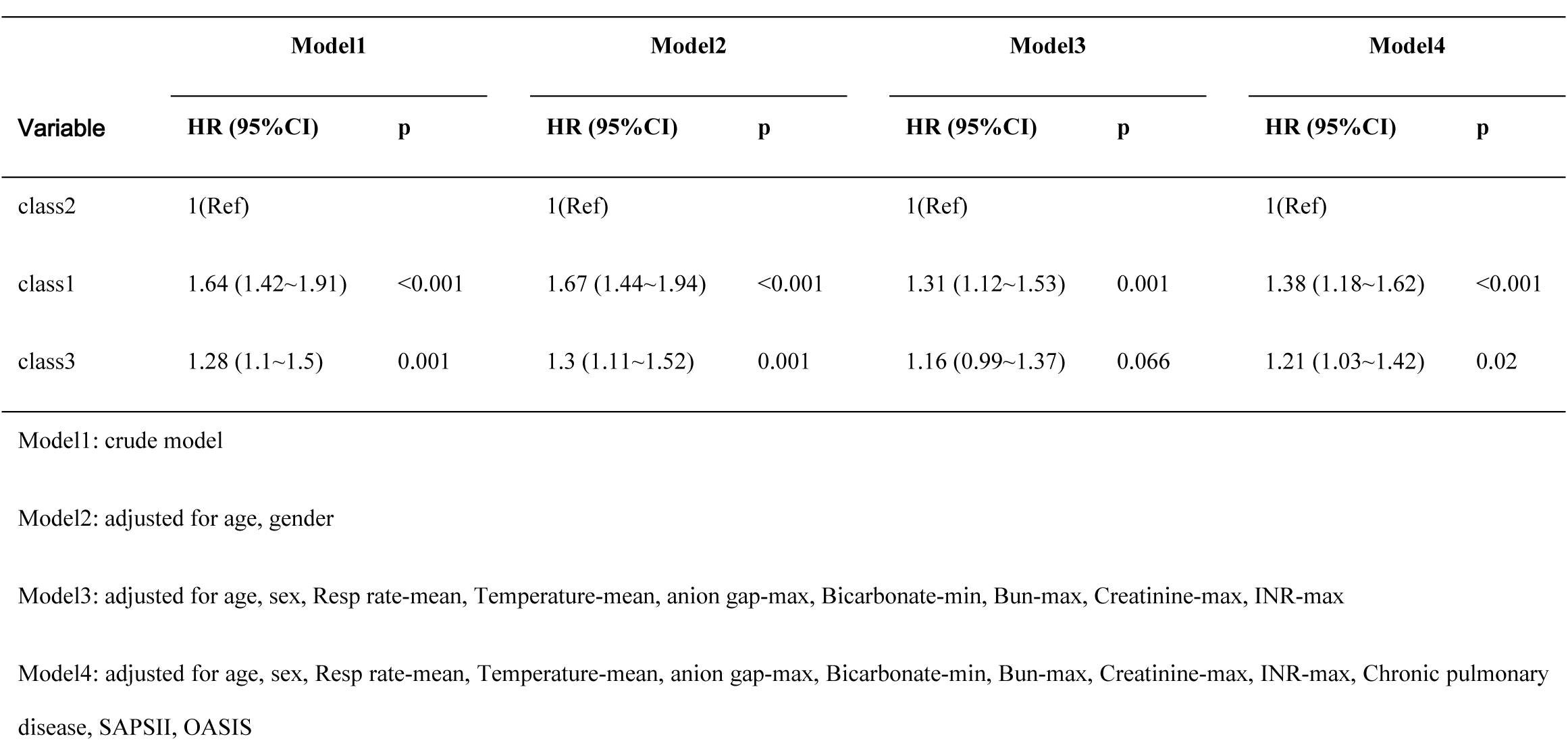
Relationship between different Hb Classes and 28-day morality in different models.

The Kaplan-Meier curves derived from the multivariate analysis indicated that varying Hb trajectories (Classes 1, 2, and 3) were significantly correlated with the 28-day survival probability (p < 0.001). Specifically, the category exhibiting the lowest Hb level (class 1) demonstrated the poorest survival rate, whereas Class 3 exhibited a survival rate higher than that of Class 1 but lower than that of Class 2, as illustrated in Fig 2.

**Fig. 2.**
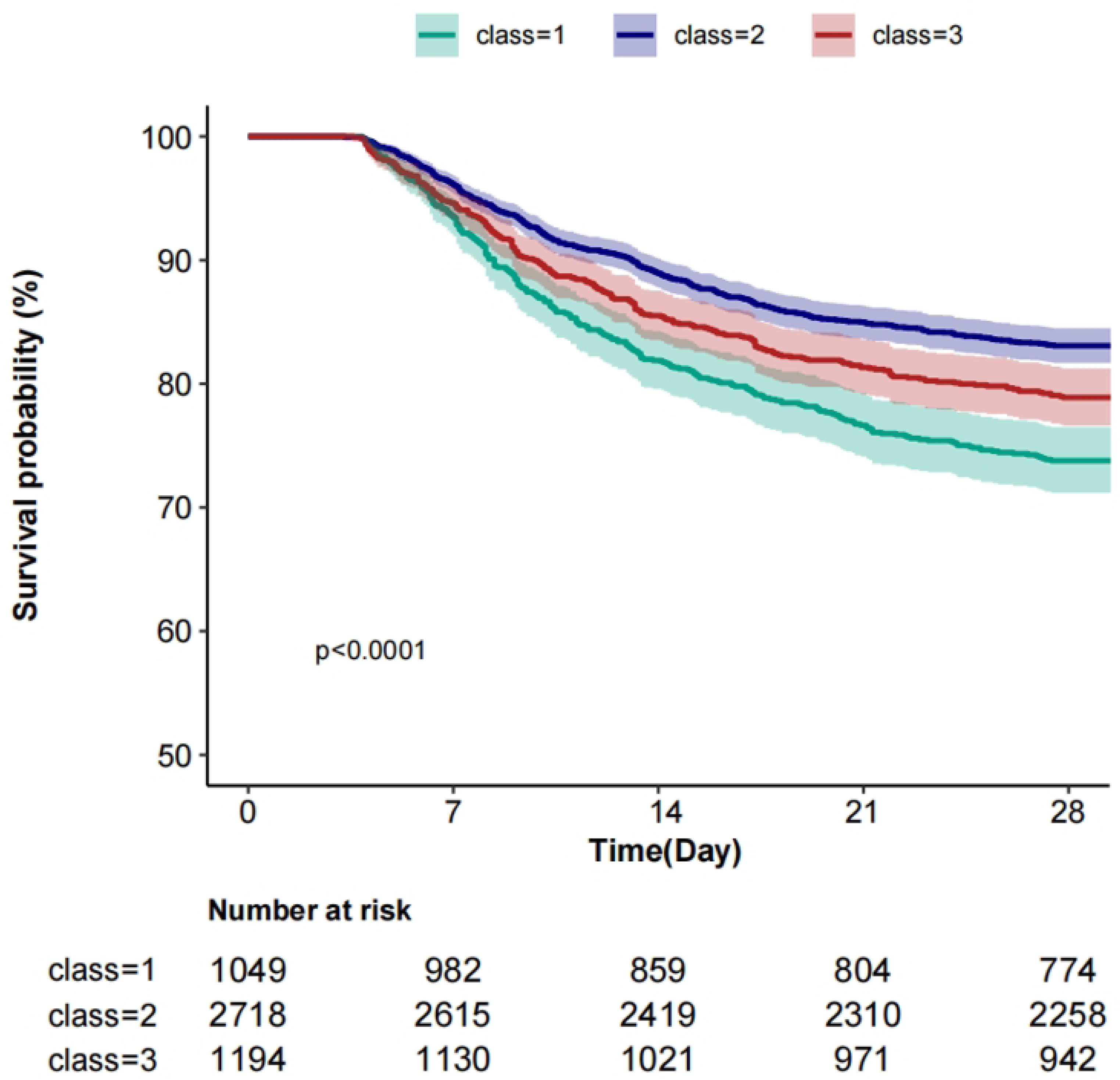
KM curves for different trajectories

### Sensitivity analysis

A subgroup analysis was conducted as part of a sensitivity analysis. S2 Fig illustrates the subgroup classifications and event occurrences among all patients, assessing the impact of various factors such as sex, age, congestive heart failure, cerebrovascular disease, diabetes, liver disease, kidney disease, malignant cancer, and the Charlson comorbidity index, SAPSII, and OASIS (with Age, Charlson comorbidity index, SAPSII, and OASIS categorized based on the median) on mortality risk, measured as the hazard ratio (HR). In all subgroup analyses for each category, the p-values were greater than 0.05, confirming the robustness of the results.

To mitigate the potential effects of unmeasured confounders, we conducted an unmeasured confounding E-value analysis. The E-value for the point estimate was 1.249, with a lower confidence interval of 1.121 and an upper confidence interval of 1.395. These relatively large E-values indicate the robustness of our findings, as illustrated in S3 Fig.

Considering the differences in Hb reference values between genders, we conducted trajectory analyses separately for males and females, as illustrated in Fig 3. Overall, males exhibited slightly higher Hb levels at various time points compared to females; however, the trajectories remained consistent across genders. In terms of Hb values at different time points, significant differences were observed between the survival and death groups for males on Day 7, Day 14, and in the minimum Hb values. For females, differences were noted between the survival and death groups on Day 1, Day 5, Day 7, Day 14, as well as in the maximum, minimum, and average values, as detailed in Table 4. The K-M curves derived from the multivariate analysis indicated that varying Hb trajectories (Classes 1, 2, and 3) were significantly correlated with the 28-day survival probability in both female and male (both p<0.05), as depicted in Fig 4.

**Fig. 3.**
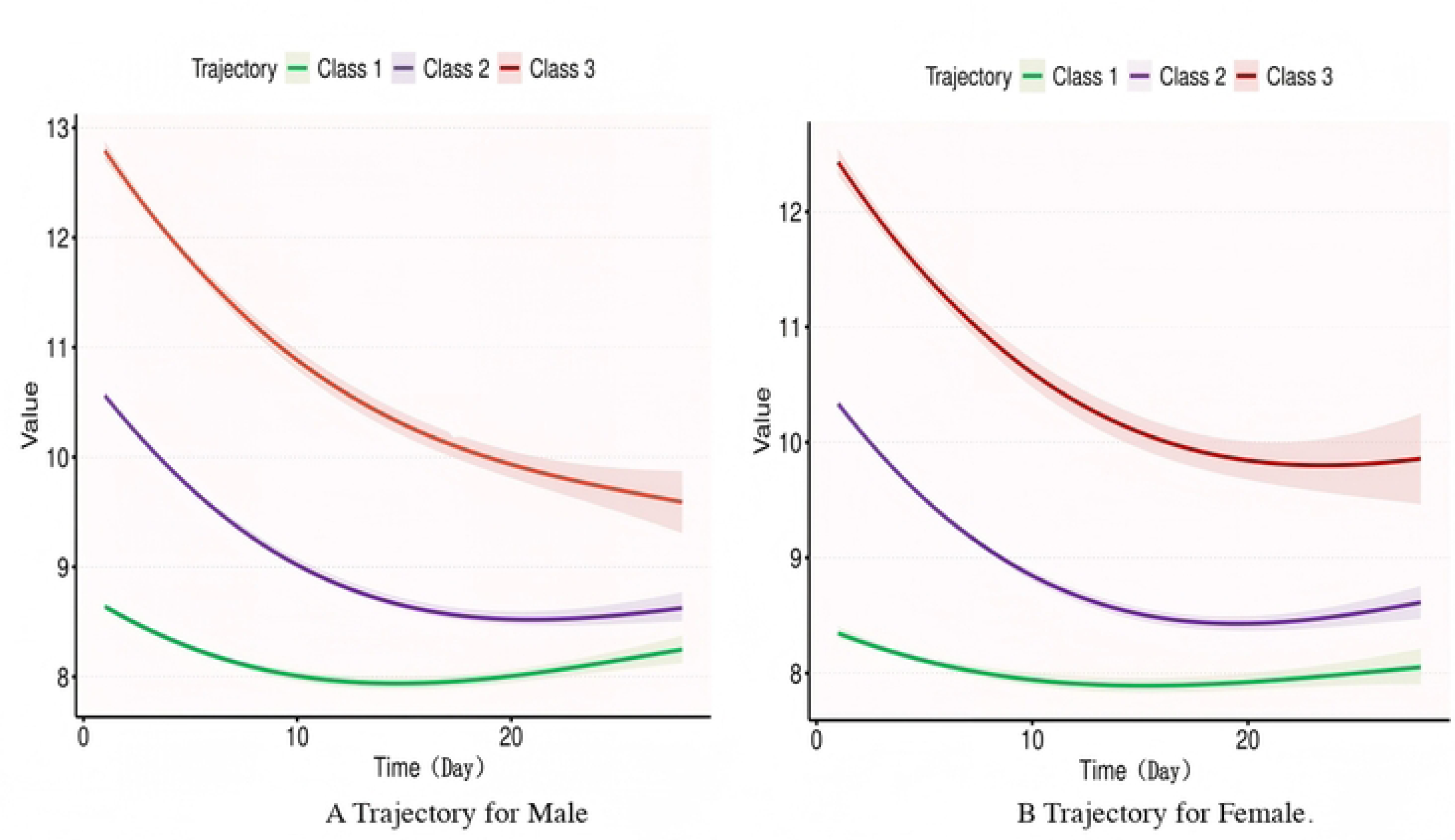

**Fig. 4.**
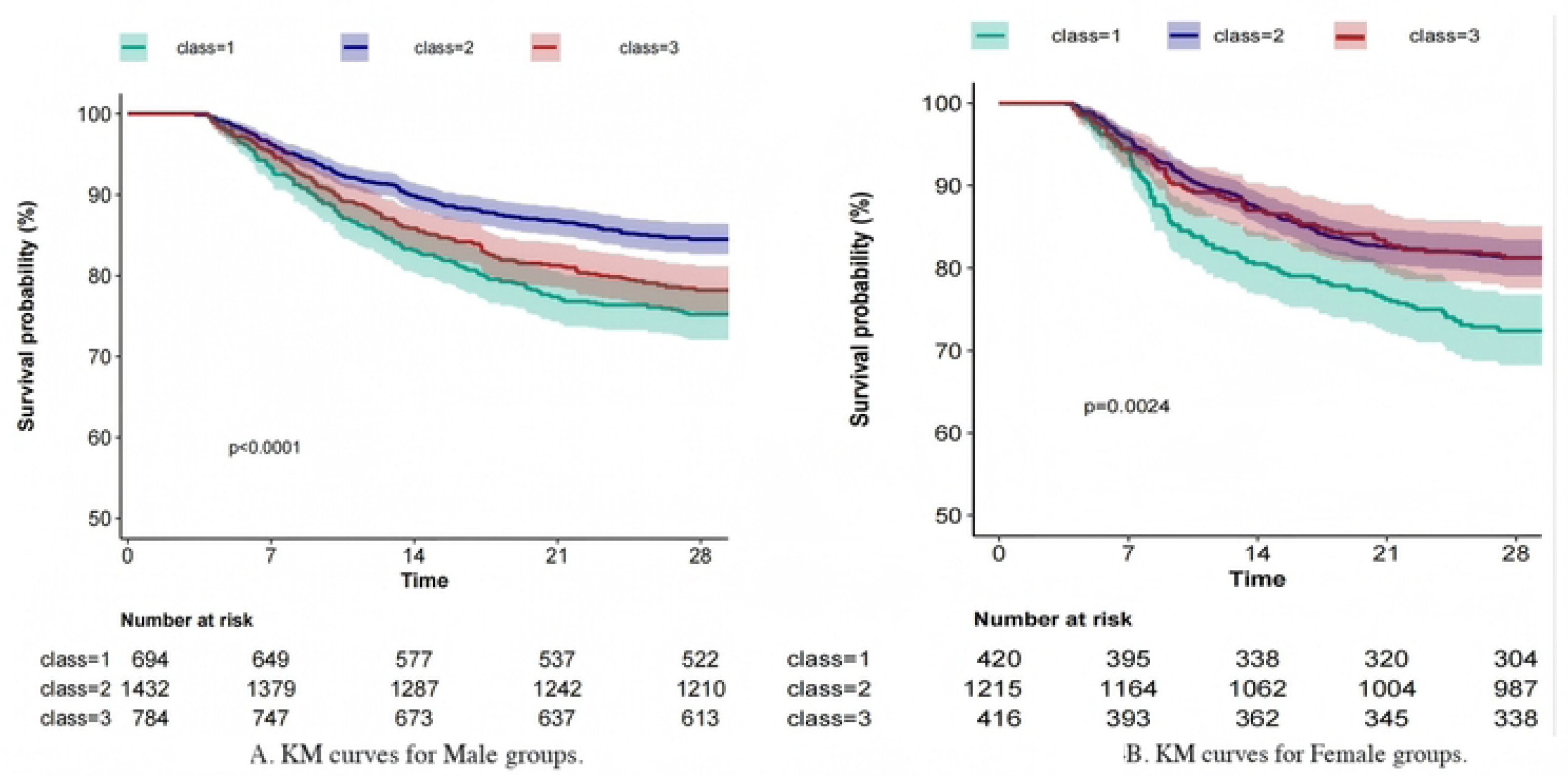

**Table 4.** Hb value Characteristics at different time points of different gender.

| Gender | Male |  |  |  | Female |  |  |  |
| --- | --- | --- | --- | --- | --- | --- | --- | --- |
|  | Total | Survival group | Death group | p | Total | Survival group | Death group | p |
| Variables | (n = 2910) | (n = 2345) | (n = 565) |  | (n = 2051) | (n = 1629) | (n = 422) |  |
| Hb Day1 | 11.0 ± 2.0 | 11.1 ± 1.9 | 11.0 ± 2.2 | 0.296 | 10.7 ± 1.8 | 10.7 ± 1.7 | 10.4 ± 1.8 | 0.001 |
| Hb Day3 | 9.8 ± 1.6 | 9.8 ± 1.6 | 9.9 ± 1.8 | 0.194 | 9.5 ± 1.5 | 9.6 ± 1.4 | 9.5 ± 1.5 | 0.254 |
| Hb Day5 | 9.8 ± 1.6 | 9.9 ± 1.6 | 9.8 ± 1.7 | 0.155 | 9.6 ± 1.5 | 9.6 ± 1.5 | 9.4 ± 1.5 | 0.001 |
| Hb Day7 | 9.8 ± 1.6 | 9.8 ± 1.6 | 9.6 ± 1.7 | 0.038 | 9.5 ± 1.5 | 9.5 ± 1.5 | 9.2 ± 1.5 | 0.001 |
| Hb Day14 | 9.2 ± 1.5 | 9.3 ± 1.5 | 9.1 ± 1.6 | 0.041 | 9.0 ± 1.3 | 9.0 ± 1.3 | 8.7 ± 1.2 | 0.002 |
| Hb Day21 | 8.8 ± 1.3 | 8.8 ± 1.3 | 8.5 ± 1.3 | 0.033 | 8.7 ± 1.1 | 8.7 ± 1.1 | 8.4 ± 1.0 | 0.098 |
| Hb Day28 | 8.7 ± 1.1 | 8.7 ± 1.1 | 8.4 ± 0.8 | 0.414 | 8.6 ± 1.1 | 8.6 ± 1.1 | 7.8 ± 1.0 | 0.087 |
| Hb-max | 11.4 ± 1.8 | 11.4 ± 1.7 | 11.3 ± 2.1 | 0.333 | 11.0 ± 1.6 | 11.1 ± 1.6 | 10.8 ± 1.7 | < 0.001 |
| Hb-min | 8.5 ± 1.7 | 8.5 ± 1.6 | 8.3 ± 1.9 | 0.027 | 8.1 ± 1.5 | 8.1 ± 1.5 | 8.0 ± 1.5 | 0.074 |
| Hb-mean | 9.8 ± 1.5 | 9.8 ± 1.4 | 9.7 ± 1.7 | 0.24 | 9.5 ± 1.3 | 9.5 ± 1.3 | 9.3 ± 1.4 | 0.012 |
| Class n (%) |  |  |  | < 0.001 |  |  |  | < 0.001 |
| 1 | 694 (23.8) | 522 (22.3) | 172 (30.4) |  | 420 (20.5) | 304 (18.7) | 116 (27.5) |  |
| 2 | 1432 (49.2) | 1210 (51.6) | 222 (39.3) |  | 1215 (59.2) | 987 (60.6) | 228 (54) |  |
| 3 | 784 (26.9) | 613 (26.1) | 171 (30.3) |  | 416 (20.3) | 338 (20.7) | 78 (18.5) |  |

The multivariate regression model indicated that the dynamic trajectory of Hb in males had a higher predictive value for 28-day survival compared to females (Table 5 and Table 6). In the unadjusted model for males, Class 1 was associated with a significantly increased risk of mortality compared to Class 2 (HR 1.70, p < 0.001), while Class 3 also exhibited an upward trend (HR 1.46, p < 0.001). After adjustments (Models 2 to 4), the risk in the Class 1 group remained significant (HR 1.21–1.67, p < 0.05). Furthermore, the risk for females in the hemoglobin Class 1 group was also significantly elevated (HR 1.38–1.59, p < 0.01), although no significant difference was observed in the risk for Class 3. These findings suggest that persistently low hemoglobin levels serve as an independent predictor of mortality across both genders.

**Table 5.**
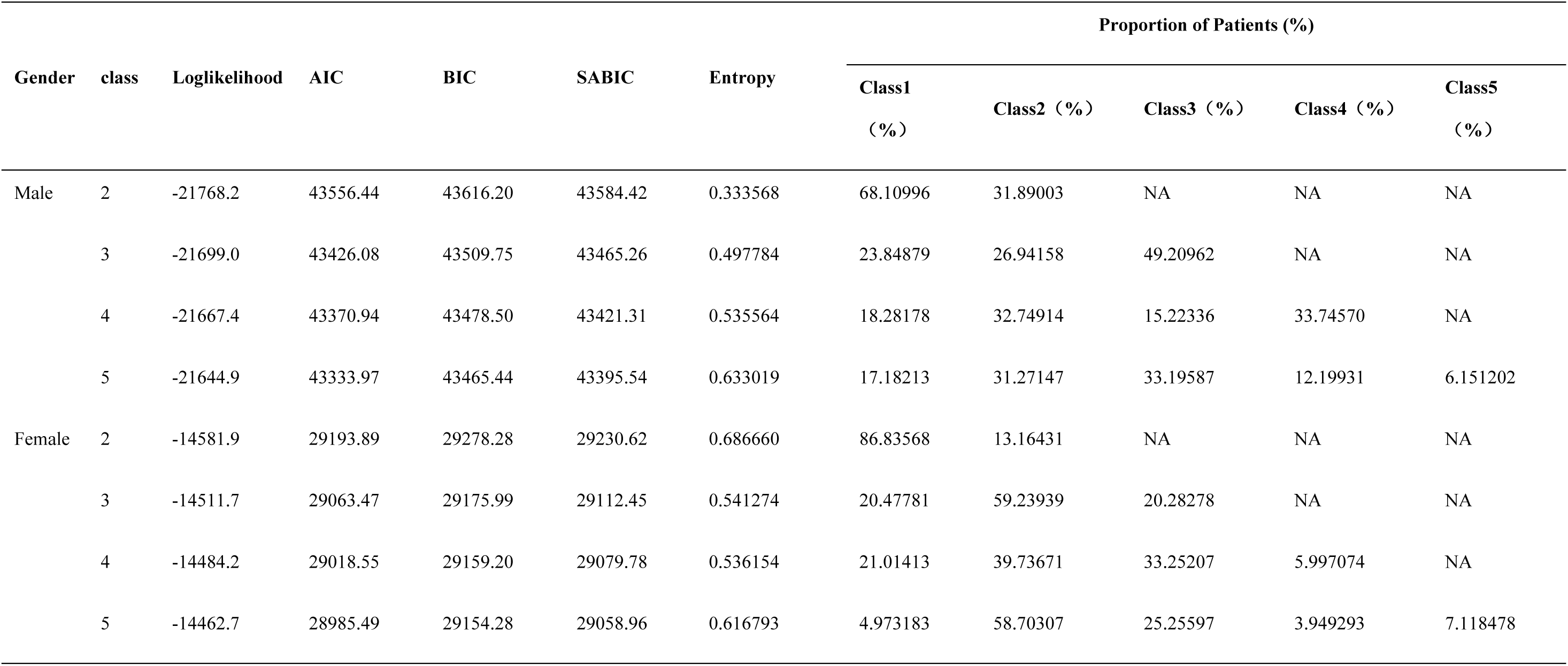
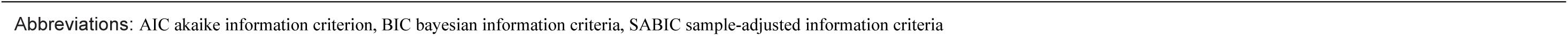
Metrics for the trajectories for Male and Female.

| Gender | class | Loglikelihood | AIC | BIC | SABIC | Entropy | Proportion of Patients (%) |  |  |  |  |
| --- | --- | --- | --- | --- | --- | --- | --- | --- | --- | --- | --- |
|  |  |  |  |  |  |  | Class1 (%) | Class2 (%) | Class3 (%) | Class4 (%) | Class5 (%) |
| Male | 2 | -21768.2 | 43556.44 | 43616.20 | 43584.42 | 0.333568 | 68.10996 | 31.89003 | NA | NA | NA |
|  | 3 | -21699.0 | 43426.08 | 43509.75 | 43465.26 | 0.497784 | 23.84879 | 26.94158 | 49.20962 | NA | NA |
|  | 4 | -21667.4 | 43370.94 | 43478.50 | 43421.31 | 0.535564 | 18.28178 | 32.74914 | 15.22336 | 33.74570 | NA |
|  | 5 | -21644.9 | 43333.97 | 43465.44 | 43395.54 | 0.633019 | 17.18213 | 31.27147 | 33.19587 | 12.19931 | 6.151202 |
| Female | 2 | -14581.9 | 29193.89 | 29278.28 | 29230.62 | 0.686660 | 86.83568 | 13.16431 | NA | NA | NA |
|  | 3 | -14511.7 | 29063.47 | 29175.99 | 29112.45 | 0.541274 | 20.47781 | 59.23939 | 20.28278 | NA | NA |
|  | 4 | -14484.2 | 29018.55 | 29159.20 | 29079.78 | 0.536154 | 21.01413 | 39.73671 | 33.25207 | 5.997074 | NA |
|  | 5 | -14462.7 | 28985.49 | 29154.28 | 29058.96 | 0.616793 | 4.973183 | 58.70307 | 25.25597 | 3.949293 | 7.118478 |

Abbreviations: AIC akaike information criterion, BIC bayesian information criteria, SABIC sample-adjusted information criteria

**Table 6.**
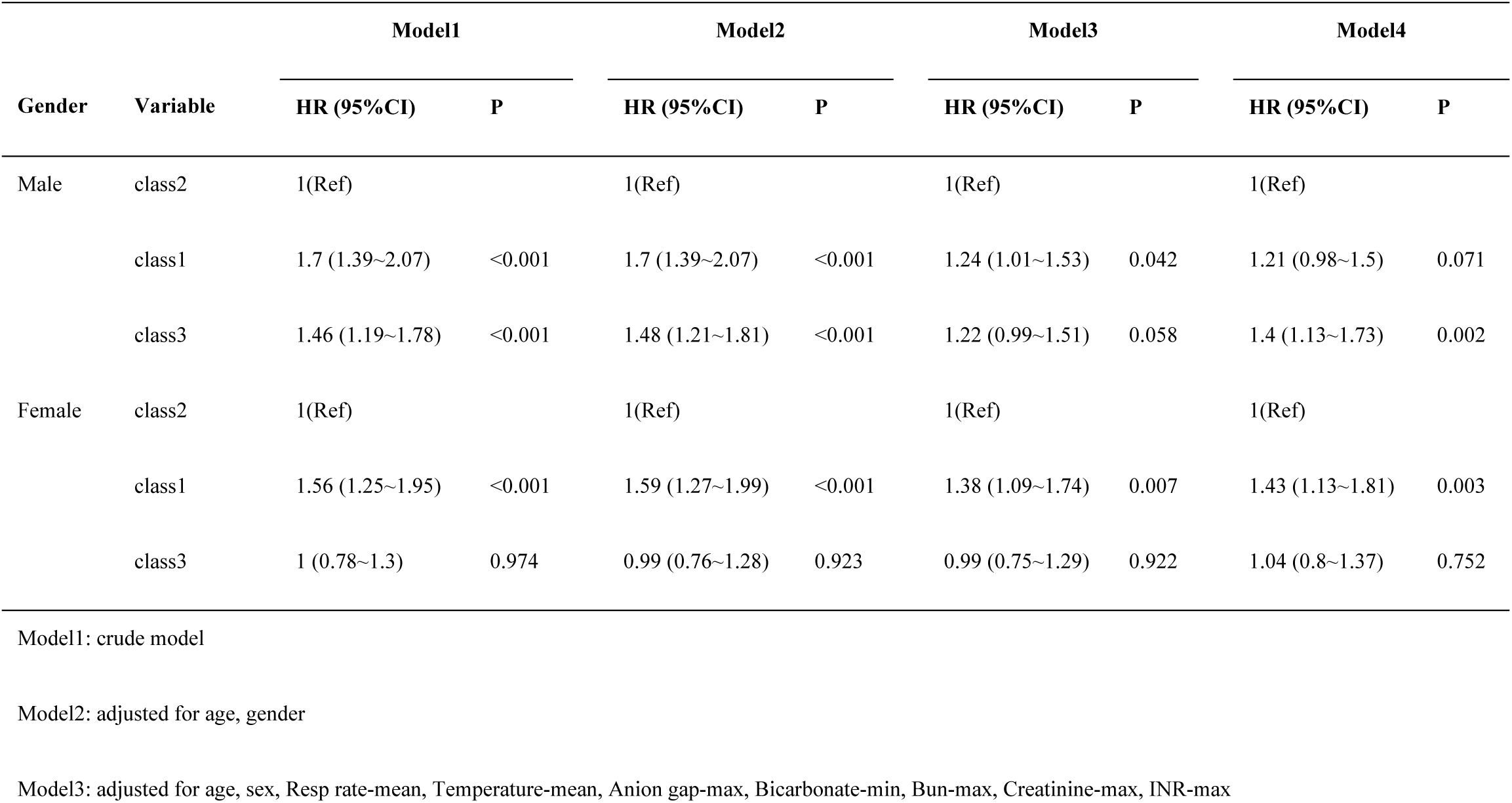
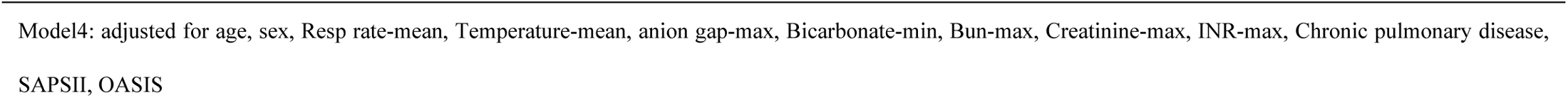
Relationship between different Hb Classes and 28-day morality in different models and Genders.

| Gender | Variable | Model1 |  | Model2 |  | Model3 |  | Model4 |  |
| --- | --- | --- | --- | --- | --- | --- | --- | --- | --- |
|  |  | HR (95%CI) | P | HR (95%CI) | P | HR (95%CI) | P | HR (95%CI) | P |
| Male | class2 | 1(Ref) |  | 1(Ref) |  | 1(Ref) |  | 1(Ref) |  |
|  | class1 | 1.7 (1.39~2.07) | <0.001 | 1.7 (1.39~2.07) | <0.001 | 1.24 (1.01~1.53) | 0.042 | 1.21 (0.98~1.5) | 0.071 |
|  | class3 | 1.46 (1.19~1.78) | <0.001 | 1.48 (1.21~1.81) | <0.001 | 1.22 (0.99~1.51) | 0.058 | 1.4 (1.13~1.73) | 0.002 |
| Female | class2 | 1(Ref) |  | 1(Ref) |  | 1(Ref) |  | 1(Ref) |  |
|  | class1 | 1.56 (1.25~1.95) | <0.001 | 1.59 (1.27~1.99) | <0.001 | 1.38 (1.09~1.74) | 0.007 | 1.43 (1.13~1.81) | 0.003 |
|  | class3 | 1 (0.78~1.3) | 0.974 | 0.99 (0.76~1.28) | 0.923 | 0.99 (0.75~1.29) | 0.922 | 1.04 (0.8~1.37) | 0.752 |
| Model1: crude model |  |  |  |  |  |  |  |  |  |
| Model2: adjusted for age, gender |  |  |  |  |  |  |  |  |  |
| Model3: adjusted for age, sex, Resp rate-mean, Temperature-mean, Anion gap-max, Bicarbonate-min, Bun-max, Creatinine-max, INR-max |  |  |  |  |  |  |  |  |  |

## Discussion

To our knowledge, this study is the first to explore the relationship between dynamic Hb value trajectories and 28-day mortality risk in elderly sepsis patients. The Cox model demonstrates that dynamic trajectories can indirectly reflect treatment effects and changes in patients’ pathophysiology compared to single Hb measurements at specific time points, thereby providing a more accurate prognostic prediction. This study indicates that trajectories of hemoglobin levels characterized by persistently low values or a rapid decrease from near-normal values are associated with an increased risk of 28-day mortality compared to trajectories that decline slowly around the median value.

For a long time, hemoglobin levels have been considered associated with disease prognosis. An initial hemoglobin level of 80 g/L or lower within the first 48 hours of ICU admission is recognized as a predictive indicator of one-year mortality in severe sepsis, underscoring the significance of early hemoglobin measurements in sepsis prognosis [22,23]. Insufficient hemoglobin can lead to inadequate tissue oxygenation, while an inappropriate increase in hemoglobin may indicate a hypercoagulable state or even thrombosis[24]. Research has demonstrated a non-linear relationship between hemoglobin levels and various diseases, including renal disease and the mortality rate during acute exacerbations of COPD[24]. Recent studies indicate that admission hemoglobin levels also exhibit a non-linear relationship with 30-day mortality in sepsis patients. Specifically, with a threshold of 7.2 g/dL, each 1 g/dL increase in hemoglobin is associated with an approximately 13% reduction in the odds ratio (OR) for 30-day mortality. This finding suggests that maintaining hemoglobin levels above 7.2 g/dL may reduce the risk of death in patients with sepsis [25].

The correlation between hemoglobin levels and 28-day mortality in elderly patients with sepsis represents an innovative study, as this demographic often presents complex pathological conditions, including anemia, malnutrition, decline in cardiopulmonary function, and other comorbidities, which complicate prognostic analysis. The average hemoglobin (Hb) concentration in the elderly population tends to gradually decrease with age[26]. Furthermore, the higher prevalence of anemia among older adults is frequently linked to various comorbidities, such as chronic kidney disease, cardiovascular disorders, and malnutrition. Additionally, elderly individuals with anemia are at an increased risk for adverse clinical outcomes, including elevated hospitalization rates, frailty, cognitive decline, and higher mortality[27]. This study reveals that hemoglobin levels in elderly patients with sepsis are comparatively lower than established normal reference values, aligning with the recognized phenomenon that critically ill elderly patients typically exhibit diminished hemoglobin levels. Notably, approximately 40% of patients experience a reduction in hemoglobin levels to 10 g/dL following hospitalization[28].

The trend of hemoglobin changes during the treatment process may indicate alterations in clinical conditions. For instance, the presence of persistently worsening anemia in a patient may suggest a deterioration in clinical status, potentially necessitating further blood transfusions to correct the anemia or enhance supportive therapy [29]. This study is distinguished by its focus on the association between hemoglobin trajectory and 28-day mortality. We collected hemoglobin data for patients upon admission and at days 1, 3, 5, 7, 14, 21, and 28 for trajectory analysis, which enhanced predictive capability. Furthermore, the results of the subgroup analysis confirmed the stability of the findings. Upon reclassifying and analyzing the trajectories by gender, we found that the trajectory classifications were largely consistent between males and females. However, the Hb values at various time points were higher in males than in females, and the predictive power of the third category’s accumulation for early mortality risk was stronger in males than in females. Additionally, we found that female, compared to male, exhibit a greater tolerance for rapid decreases in hemoglobin levels and a relatively lower risk of adverse outcomes.

Early studies indicate gender differences in anemia tolerance. Males exhibit higher mortality rates at lower nadir hemoglobin (Hb) levels, particularly at 6.0 g/dL or less, whereas females do not show a corresponding risk increase. Adjusted for confounders, inpatient mortality is significantly higher in males under similar conditions, suggesting females better tolerate lower Hb levels[30]. The differences in anemia tolerance between genders may arise from distinct physiological bases and response mechanisms. Although females typically present with lower baseline hemoglobin levels, they have biologically adapted to low hemoglobin states through menstrual and pregnancy-related blood loss, which enhances their recovery capacity during episodes of anemia, thereby maintaining normal physiological functions. Furthermore, estrogen and other sex hormones play a crucial role in regulating erythropoiesis and hemodynamics, potentially exacerbating the differences in anemia responses between gender[30]. Collectively, these factors elucidate the advantages that females possess in terms of anemia tolerance. In this study, the differences in the trajectory of decline in predictive ability for mortality risk correspond with the variations in anemia tolerance between men and women.

Our study possesses several strengths. Firstly, it emphasizes the 28-day mortality rates among elderly sepsis patients, a demographic typically characterized by more severe disease states, a higher burden of comorbidities, and an increased need for clinical intervention. Secondly, this research leverages public databases and employs repeated hemoglobin measurements from the first month of ICU hospitalization to establish dynamic trajectory patterns. Furthermore, it conducts proportional hazards testing to explore potential time cutoff points within the study duration. Finally, the JLCM method serves as a form of unsupervised trajectory analysis, facilitating the identification of valuable insights. Additionally, we conducted a series of sensitivity analyses, including the Schoenfeld residual test, subgroup analysis, and E-value testing, to ensure the robustness of our study results.

This study has several inherent limitations. The analytical data were obtained from a public database of a single research institution, which restricted the extraction and joint analysis of imaging features, treatment protocols, and nursing measures, as well as the monitoring of additional therapeutic outcomes. The E-value is utilized as a method for sensitivity analyses to clarify the extent to which the study results may be influenced by unmeasured confounding factors. Nevertheless, the relatively large E-values bolster the reliability of the findings.

## Conclusion

In summary, dynamic analysis of hemoglobin trajectory provides critical prognostic information regarding mortality in elderly patients with sepsis. Effective monitoring and management of hemoglobin may be a key strategy for improving clinical outcomes in this high-risk group, addressing the dual challenges of sepsis and anemia. Further research aimed at optimizing treatment approaches based on these dynamic trajectories could ultimately enhance clinical prognoses for this specific population.

## Author contributions

**Conceptualization:** Yan Zeng.

**Data curation:** Jingwei Zhang.

**Formal analysis:** Caoyi Liu.

**Funding acquisition:** Jingwei Zhang.

**Investigation:** Caoyi Liu.

**Methodology:** Jingwei Zhang.

**Software:** Yan Zeng.

**Supervision:** Jun Wang.

**Validation:** Jun Wang.

**Visualization:** Yan Zeng.

**Writing** – original draft: Yan Zeng.

**Writing** – review & editing: Jingwei Zhang.

## Data accessibility

Datasets were sourced from the MIMIC-IV database (version 3.0), which is available at https://physionet.org/content/mimiciv/3.0/. Authorization codes 13813672 for Zhang facilitated access.

## Ethical considerations

Compliant with local and institutional regulations, this research on human subjects did not necessitate ethical approval or subsequent reviews. Additionally, obtaining written informed consent was not required by either national laws or institutional policies.

## Acknowledgements

We express our profound gratitude to the Massachusetts Institute of Technology and the Beth Israel Deaconess Medical Center for their invaluable contribution to the MIMIC project.

## Supporting information

S1 Fig. Flowchart of study population.

S2 Fig.Forest plot of subgroup analyses.

S3 Fig. E value bias plot.

S1 Table. Clinical Characteristics of Different Hemoglobin Trajectories.

S2 Table. Univariate Cox Regression Analysis Results for 28-Day Mortality.

S3 Table. The results of covariate selection.

